# Attitudes and Perceptions of Generative Artificial Intelligence Chatbots in the Scientific Process of Traditional, Complementary, and Integrative Medicine Research: A Large-Scale, International Cross-Sectional Survey

**DOI:** 10.64898/2026.04.13.26350612

**Authors:** Jeremy Y. Ng, Jamie Tan, Niveen Syed, Karthik Adapa, Prashant Kumar Gupta, Shao Li, Darshan Mehta, Melinda Ring, Manisha Shridhar, Joao Paulo Souza, Tetsuhiro Yoshino, Myeong Soo Lee, Holger Cramer

## Abstract

**Background:** Generative artificial intelligence (GenAI) chatbots have shown utility in assisting with various research tasks. Traditional, complementary, and integrative medicine (TCIM) is a patient-centric approach that emphasizes holistic well-being. The integration of TCIM and GenAI presents numerous key opportunities. However, TCIM researchers’ attitudes toward GenAI tools remain less understood. This large-scale, international cross-sectional survey aimed to elucidate the attitudes and perceptions of TCIM researchers regarding the use of GenAI chatbots in the scientific process.

**Methods:** A search strategy in Ovid MEDLINE identified corresponding authors who were TCIM researchers. Eligible authors were invited to complete an anonymous online survey administered via SurveyMonkey. The survey included questions on socio-demographic characteristics, familiarity with GenAI chatbots, and perceived benefits and challenges of using GenAI chatbots. Results were analysed using descriptive statistics and thematic content analysis.

**Results:** The survey received 716 responses. Most respondents reported familiarity with GenAI chatbots (58.08%) and viewed them as very important to the future of scientific research (54.37%). The most acknowledged benefits included workload reduction (74.07%) and increased efficiency in data analysis/experimentation (71.14%). The most frequently reported challenges involved bias, errors, and limitations. More than half of the respondents (57.02%) expressed a need for training to use GenAI chatbots in the scientific process, alongside an interest in receiving training (72.07%). However, 43.67% indicated that their institutions did not offer these programs.

**Conclusion:** By developing a deeper understanding of TCIM researchers’ perspectives, future AI applications in this field can be more informed, and guide future policies and collaboration among researchers.

## Background

### Introduction to Artificial Intelligence in Scientific Research

Artificial intelligence (AI) has become increasingly integrated into the scientific process, providing tools to enhance efficiency, accuracy, and objectivity in various research activities [1,2]. Conventional AI systems, such as machine learning models, have aided research tasks including statistical modelling, image analysis, and data classification [3]. More recently, generative AI (GenAI) tools, a subset of AI encompassing large language models like OpenAI’s ChatGPT and Google Gemini, hold particular promise for supporting researchers by streamlining workflows, fostering collaboration, and democratizing access to advanced computational tools [4]. These technologies can potentially enhance productivity and innovation while ensuring rigour and reproducibility in the scientific process [5]. GenAI chatbots can streamline tasks such as literature searches and reviews, analyse and interpret large datasets, and aid in drafting manuscripts [6,7]. They can also enhance the readability of scientific articles among non-native speakers, potentially decreasing language barriers in research [8]. In contrast, concerns have been raised regarding integrating GenAI tools into scientific research. These include the potential for factual inaccuracies, lack of transparency in decision-making, potential amplification of biases, data privacy issues, and questions surrounding authorship and accountability [4]. Overreliance on such tools may also negatively affect critical thinking and writing skills, which could have greater implications for novice researchers [3]. Recognizing the opportunities and limitations of GenAI is vital for its responsible and equitable integration into research. These considerations have implications for clinical care, as research informs evidence-based outcomes and patient care.

### The Role of Artificial Intelligence in Enhancing Traditional, Complementary, and Integrative Medicine Research Methodology and Accessibility

Traditional, complementary, and integrative medicine (TCIM) encompasses diverse practices, including herbal medicine, mind-body practices (e.g., yoga, meditation), Ayurveda, and homeopathy, which often fall outside the scope of conventional biomedical frameworks [9]. Traditional medicine refers to the collective knowledge and practices derived from various cultures, encompassing long-established beliefs, skills, and methods for health promotion, disease prevention, diagnosis, and treatment of mental and physical illnesses [10]. Complementary medicine is defined by the World Health Organization (WHO) as “additional healthcare practices that are not part of a country’s mainstream medicine” [11]. Comparatively, integrative medicine is defined by the WHO as an “interdisciplinary and evidence-based approach to health and well-being by using a combination of biomedical and traditional and/or complementary medical knowledge, skills and practices”, emphasizing holistic well-being [11]. In contrast, the National Center for Complementary and Integrative Health (NCCIH) defines complementary approaches as those used together with conventional medicine, and integrative health as combining conventional and complementary approaches in a way to emphasize holistic, patient-centred care [12,13]. As TCIM research is deeply intertwined with culturally grounded knowledge systems and diverse epistemologies, understanding TCIM-specific attitudes toward GenAI is essential to ensure these technologies are integrated in ways that respect TCIM-specific methodologies and avoid the misappropriation of traditional knowledge [14]. Conducting rigorous research in TCIM can be particularly complex due to the interdisciplinary nature of the field, its reliance on diverse methodologies, and challenges associated with validating treatments [14,15]. GenAI chatbots can provide unique support to TCIM researchers by assisting with tasks such as synthesizing complex information, designing robust methodologies, and improving the clarity and coherence of research outputs [3]. These tools may also contribute to the equitable advancement of TCIM research by bridging gaps in resources and expertise among researchers from different regions and disciplines. Given the rising number of articles being published in TCIM [16] and the increasing prevalence of patients incorporating TCIM into their healthcare [17], GenAI tools may play an increasingly important role in supporting evidence-based practices and helping researchers monitor developments in the TCIM field.

### Gap in Knowledge

Despite the transformative potential of GenAI in research [18], little is known about how TCIM researchers perceive the role of GenAI chatbots in the scientific process. There is a notable lack of empirical data on their attitudes toward these tools, including their perceived benefits, limitations, and ethical concerns. Understanding these attitudes and perceptions is crucial to informing the responsible development and implementation of GenAI chatbots in ways that align with the unique needs and values of the TCIM research community. Additionally, addressing concerns about transparency, accountability, and biases is essential to fostering trust and acceptance among researchers [19,20].

### Relevance and Impact

Investigating the attitudes and perceptions of TCIM researchers toward GenAI chatbots in the scientific process is both timely and significant. By automating routine tasks, providing intelligent recommendations, and enhancing data interpretation, GenAI chatbots can potentially reduce researchers’ workload and improve the overall efficiency and quality of scientific inquiry [18]. Furthermore, it is already known that these tools may help researchers address complex challenges in TCIM research related to fostering interdisciplinary collaboration by synthesizing information across multiple disciplines, translating terminology, facilitating communication between researchers from diverse backgrounds and specializations, and enhancing methodological rigour [21]. For example, AI-powered clinical decision support systems integrate data from multiple sources to provide evidence-based recommendations, facilitating communication among healthcare professionals and improving patient outcomes [18,21]. Exploring ethical considerations, such as data security and algorithmic bias, will be critical to ensuring the responsible adoption of GenAI chatbots in this field [18,21]. Intellectual property remains a foundational element that enables access to traditional knowledge for research and represents a core ethical concern requiring careful navigation to prevent misuse of GenAI chatbots [22]. Insights from this study may help guide the development and integration of GenAI tools that align with the goals of TCIM researchers and support the advancement of evidence-based TCIM practices.

In summary, this study aims to fill an existing knowledge gap by exploring the attitudes and perceptions of TCIM researchers regarding the use of GenAI chatbots in the scientific process. The findings may offer useful insights into how these tools could be responsibly implemented to support TCIM research, with potential implications for the broader scientific community.

## Methods

### Approach and Open Science Statement

This study used a cross-sectional, web-based survey design targeting TCIM researchers identified as corresponding authors of subject-specific MEDLINE-indexed articles. This study’s protocol was registered on the Open Science Framework (OSF) [23]. Before any data collection, ethics approval was obtained from the University Hospital Tubingen Research Ethics Board (REB #: 079/2025BO2). Study materials and data have been shared on OSF and can be found at https://doi.org/10.17605/OSF.IO/VEZRF. After all data collection and analysis, the findings were incorporated into a manuscript and posted as a preprint before submission to a peer-reviewed journal. No personal data was collected or processed in this study, and IP addresses were not stored. Consent for data collection, storage, use, and sharing was obtained through a consent form. Participants could withdraw from the online survey at any time.

Potential sources of bias were addressed by combining careful sampling and recruitment, validated survey administration and data collection processes, independent and inductive qualitative coding, and transparent reporting with pre-registration and open data. The manuscript was reported in accordance with the Strengthening the Reporting of Observational Studies in Epidemiology (STROBE) guidelines for cross-sectional studies and the Checklist for Reporting Results of Internet E-Surveys (CHERRIES) standards [24, 25].

### Sampling Framework

We conducted a search on Ovid MEDLINE to identify relevant articles published between December 15, 2014 and December 15, 2024. The search targeted publications related to a broad range of TCIM disciplines. The search strategy (see https://osf.io/68rjc) combined MeSH terms and keywords for key TCIM domains, including acupressure, acupuncture, chiropractic, complementary therapies, Chinese herbal medicine, electroacupuncture, herbal medicine, integrative medicine, applied kinesiology, osteopathic manipulation, Ayurvedic medicine, traditional Chinese medicine, mind-body therapies, naturopathy, phytotherapy, medicinal plants, and yoga. Based on this search strategy, it was assumed that a significant proportion of the corresponding authors of these articles were TCIM researchers. Non-research articles, such as editorials, opinions, and commentaries, were excluded through a combination of search strategy lines. PubMed Identifier (PMID) numbers from the resulting articles were exported from Ovid as .csv files and processed using an R script (based on the 2019 easyPubMed and RedEye packages) to extract author names, affiliated institutions, and email addresses [26,27]. Once the PMIDs were imported to R, the corresponding author email extraction was finalized, with the data subsequently cleaned for errors or duplicates before survey distribution. Additionally, extracted email addresses were manually checked for truncations and verified using a hex recorder developed in PyCharm before survey distribution. Authors who met the inclusion criteria as outlined in the sampling framework were invited to participate in an anonymous online survey. Details about the process for retrieving authors’ names and email addresses can be found at https://osf.io/vezrf/files/68rjc.

### Participant Recruitment

Only researchers identified through the sampling framework were invited to participate in the study. These participants were contacted via email and invited to complete the survey voluntarily and anonymously. Emails were sent via SurveyMonkey [28], including details about the study objectives and a link to an informed consent form. The consent form provided information on the estimated time required to complete the survey, how and where data would be stored and for how long, the identity of the investigator, and the purpose of the study (see https://osf.io/vezrf/files/v64eg). Participants were required to provide consent before accessing the online survey questions. Duplicate entries of name and email address pairs were removed from the sample. A response rate of 3-7% was anticipated based on prior research conducted by JYN on related topics [6,29,30]. The survey was open from 18 August 2025 to 22 September 2025. Participants had five weeks to complete the survey, with reminder emails sent one, two, and three weeks after the initial invitation. A two-week final-response window followed the final reminder email. Copies of the initial and follow-up email invitations can be found at https://osf.io/vezrf/files/u4akb. Participation was entirely voluntary, with no financial compensation provided.

### Survey Design

The closed survey was developed using SurveyMonkey software [28]. Researchers received an email invitation containing a link to the survey on SurveyMonkey [28], which began with an implied consent agreement that participants had to agree to before viewing the survey questions. The survey consisted of multiple-choice questions, beginning with demographic questions. The demographic section included questions adapted from previous studies on TCIM [29,30]. Following the demographics section, participants answered questions regarding their awareness, attitudes, and use of GenAI chatbots at different stages of the scientific process, including study design, data analysis, manuscript preparation, and peer communication. These AI-related questions were adapted from a recent study on AI use in research workflows [6]. The questions were distributed across eight pages, with the number of questions varying per page. The content, number, and order of questions remained consistent for all participants. Respondents were able to review and change their responses before submission, as well as skip any questions they did not wish to answer. The survey was reviewed by the authors (JYN, KA, PKG, SL, DM, MR, MS, JPS, TY, MSL, and HG), all of whom are experts in TCIM and/or AI, before circulation. The survey was expected to take approximately 15 minutes to complete. A complete version of the survey is provided at https://osf.io/vezrf/files/4unvk.

### Data Analysis

This study did not include formal hypotheses. Data from participants who did not complete the survey in full were still considered. Descriptive statistics on quantitative data, such as frequencies and percentages, were automatically collected and categorized using SurveyMonkey [28]. Subgroup differences were examined by stratifying responses across key demographic variables (i.e., age, sex, location, career stage) [28]. In addition, analyses were stratified by whether participants identified TCIM as their primary or secondary research area, allowing for comparisons based on their depth of engagement in TCIM research. If a significant number of valuable qualitative responses were collected, a thematic content analysis was performed. Two members of the research team independently coded the qualitative data and reached consensus on the coding framework. Using an inductive approach, the coding categories were developed directly from the data, with careful attention to ensuring that each category was clear, distinct, and accurately reflected participants’ responses [31,32].

## Results

After deduplication of the list of researchers’ emails, survey invitations were sent to 59,045 corresponding authors. Of these, 22,071 (37.38%) opened the survey invitation, while 31,270 (52.96%) did not, perhaps due to outdated contact information, email overload, or vacation/retirement reasons. Among the authors who opened the invitation, 716 submitted at least one survey response, corresponding to an open invitation response rate of 3.24% and an overall response rate of 1.21%. A total of 642 (89.66%) respondents met the eligibility criteria. Of these, 566 respondents completed the survey in full (88.00%), and those who did not may have been limited by eligibility constraints or time restrictions. The raw survey data is available online (see https://osf.io/vezrf/files/642bs). Because not all respondents answered every survey item, the total number of responses varied across questions. Incomplete survey responses were also analysed. Additionally, responses were stratified by age, sex, location of employment, career stage, and TCIM as a primary or secondary research focus through SurveyMonkey [28], and can be found online (see https://osf.io/vezrf/files/rujdq). The survey took an average of 11 minutes and 43 seconds to complete.

Demographic data can be found in **Table 1**. Most respondents were male (54.34%, 338/622), while less than half were female (43.41%, 270/622). A majority of respondents were aged 35-44 (29.86%, 186/623), followed by 45-54 (27.45%, 171/623) and 55-64 (19.10%, 119/623). Most respondents did not identify as being part of a visible minority group (76.38%, 474/618). Respondents primarily worked in South-East Asia (31.65%, 194/613), Europe (21.53%, 132/613), and the Americas (20.23%, 124/613). Given the option to choose more than one role, most identified as faculty members at university or academic institutions (52.64%, 389/739 responses), academic research staff (18.13%, 134/739 responses), or clinician researchers (15.16%, 112/739 responses). The largest proportion of respondents were senior researchers (65.67%, 375/571) and had published less than 100 research articles (72.63%, 398/548). For reporting purposes, the continuous variable ‘number of articles published’ was categorized into increments of 100.

**Table 1:** TCIM Researchers’ Demographics.

| **Sex** | **Responses** | **Percentage** |
| --- | --- | --- |
| Female | 270 | 43.41% |
| Male | 338 | 54.34% |
| Intersex | 1 | 0.16% |
| Prefer not to say | 12 | 1.93% |
| Prefer to self-describe | 1 | 0.16% |
| TOTAL | 622 | 100.00% |
| **Age** | **Responses** | **Percentage** |
| Under 18 | 0 | 0.00% |
| 18-24 | 6 | 0.96% |
| 25-34 | 56 | 8.99% |
| 35-44 | 186 | 29.86% |
| 45-54 | 171 | 27.45% |
| 55-64 | 119 | 19.10% |
| 65-74 | 61 | 9.79% |
| 75 or older | 19 | 3.05% |
| Prefer not to say | 5 | 0.80% |
| TOTAL | 623 | 100.00% |
| **Minority Group Status** | **Responses** | **Percentage** |
| Yes | 100 | 16.18% |
| No | 472 | 76.38% |
| Prefer not to say | 46 | 7.44% |
| TOTAL | 618 | 100.00% |
| **Country of Employment** | **Responses** | **Percentage** |
| Africa | 77 | 12.56% |
| Americas | 124 | 20.23% |
| Eastern Mediterranean | 31 | 5.06% |
| Europe | 132 | 21.53% |
| South-East Asia | 194 | 31.65% |
| Western Pacific | 55 | 8.97% |
| TOTAL | 613 | 100.00% |
| **Current Position (Select all that apply)** | **Responses** | **Percentage** |
| Faculty member at a university/academic institution | 389 | 52.64% |
| Academic research staff (e.g., research coordinator at a university/academic institution) | 134 | 18.13% |
| Government researcher | 23 | 3.11% |
| Clinician researcher | 112 | 15.16% |
| Pharmaceutical industry/company employee | 11 | 1.79% |
| Scholarly communications employee (e.g., scholarly journals/publishing, medical writing) | 6 | 0.81% |
| Consultant (e.g., own or employed by research consulting firm) | 21 | 2.84% |
| Third sector (e.g., NGO, non-profit) | 9 | 1.22% |
| Other (please specify) | 34 | 4.60% |
| TOTAL | 739 <sup>a</sup> | 100.00% |
| **Career Stage** | **Responses** | **Percentage** |
| Early career researcher (<5 years) | 76 | 13.31% |
| Mid-career researcher (5-10 years) | 120 | 21.02% |
| Senior researcher (>10 years) | 375 | 65.67% |
| TOTAL | 571 | 100.00% |
| **Primary Research Area** | **Responses** | **Percentage** |
| Clinical research | 225 | 37.25% |
| Preclinical research – in vivo | 91 | 15.07% |
| Preclinical research – in vitro | 86 | 14.24% |
| Health systems research | 23 | 3.81% |
| Health services research | 38 | 6.29% |
| Methods research | 47 | 7.78% |
| Epidemiological research | 16 | 2.65% |
| Other (please specify) | 78 | 12.91% |
| TOTAL | 604 | 100.00% |
| **Number of Published Research Articles** | **Responses** | **Percentage** |
| Under 100 | 398 | 72.63% |
| 100-199 | 103 | 18.80% |
| 200-299 | 27 | 4.93% |
| 300-399 | 12 | 2.19% |
| 400 or more | 8 | 1.46% |
| TOTAL | 548 | 100.00% |
| **TCIM Research Focus** | **Responses** | **Percentage** |
| Primary (i.e., most of my research is about TCIM) | 343 | 59.76% |
| Secondary (i.e., most of my research is not about TCIM) | 231 | 40.24% |
| TOTAL | 574 | 100.00% |
| **Most TCIM Research Conducted in:** | **Responses** | **Percentage** |
| Mind-body therapies (e.g., meditation, biofeedback, hypnosis, yoga, tai chi, imagery, creative outlets) | 84 | 14.89% |
| Biologically based practices (e.g., vitamins and dietary supplements, botanicals, special foods and diets) | 185 | 32.80% |
| Manipulative and body-based practices (massage, chiropractic therapy, reflexology) | 52 | 9.22% |
| Biofield therapy (e.g., Reiki, therapeutic touch) | 10 | 1.77% |
| Whole medical systems (e.g., Ayurvedic medicine, East Asian medicine, acupuncture, homeopathy, naturopathic medicine) | 137 | 24.29% |
| Other, please specify (e.g., methodology, | 96 | 17.02% |
| CAIM professions, etc.) |  |  |
| TOTAL | 564 | 100.00% |

<sup>a</sup> Participants could choose more than one option.

Respondents’ familiarity with GenAI chatbots and views on using GenAI chatbots in the scientific process are shown in **Table 2**. Notably, 58.08% (327/563) were familiar with the concept of GenAI chatbots. In a multiple-response question (select all that apply), ChatGPT was the most frequently reported chatbot used in the past (46.10%, 461/1000 responses). It was also the most used GenAI chatbot for purposes relating to the scientific process (44.84%, 352/785 responses), while 18.73% (147/785 responses) reported never using a GenAI chatbot in this context. More than half of respondents reported being likely (39.04%, 219/561) or very likely (37.08%, 208/561) to use GenAI chatbots for their future research. Similarly, over half of respondents felt that GenAI chatbots would be very important (54.37%, 280/515) or important (34.37%, 177/515) to future scientific research. A significant number of respondents viewed GenAI chatbots as having a positive impact (48.45%, 250/516) on the future of scientific research, with a quarter (25.19%, 130/516) expressing very positive views toward their potential contributions. In contrast, only 8.72% (45/516) felt that GenAI chatbots would negatively affect future scientific research.

**Table 2:** TCIM Researchers’ Familiarity, Use, Training, and Perceptions of GenAI in the Scientific Process.

| **How familiar are you with GenAI chatbots?** | **Responses** | **Percentage** |
| --- | --- | --- |
| Very familiar | 90 | 15.99% |
| Familiar | 327 | 58.08% |
| Unfamiliar | 93 | 16.52% |
| Very unfamiliar | 35 | 6.22% |
| I'm not sure | 18 | 3.20% |
| TOTAL | 563 | 100.00% |
| **Which GenAI chatbots have you used in the past for any purpose? (Select all that apply)** | **Responses** | **Percentage** |
| ChatGPT | 461 | 46.10% |
| Google Gemini (formerly Bard) | 178 | 17.80% |
| Meta AI | 48 | 4.80% |
| Microsoft Copilot (formerly Bing Chat) | 181 | 18.10% |
| I have never used a GenAI chatbot | 56 | 5.60% |
| Other (please specify) | 76 | 7.60% |
| TOTAL | 1000 <sup>a</sup> | 100.00% |
| **Which GenAI chatbots have you used in the past for scientific processes? (Select all that apply)** | **Responses** | **Percentage** |
| ChatGPT | 352 | 44.84% |
| Google Gemini (formerly Bard) | 104 | 13.25% |
| Meta AI | 22 | 2.80% |
| Microsoft Copilot (formerly Bing Chat) | 87 | 11.08% |
| I have never used a GenAI chatbot for purposes relating to scientific processes | 147 | 18.73% |
| Other (please specify) | 73 | 9.30% |
| TOTAL | 785 <sup>a</sup> | 100.00% |
| **How likely are you to use GenAI chatbots for research in the future?** | **Responses** | **Percentage** |
| Very likely | 208 | 37.08% |
| Likely | 219 | 39.04% |
| Unlikely | 43 | 7.66% |
| Very unlikely | 35 | 6.24% |
| I am not sure | 56 | 9.98% |
| TOTAL | 561 | 100.00% |
| **Has your research institution provided any education/training on appropriate GenAI use in scientific processes?** | **Responses** | **Percentage** |
| Yes, and I have taken it | 92 | 16.40% |
| Yes, but I have not taken it | 115 | 20.50% |
| No | 245 | 43.67% |
| I am not sure | 109 | 19.43% |
| TOTAL | 561 | 100.00% |

| **Has your research institution implemented any policies surrounding the use of GenAI chatbots in scientific processes?** | **Responses** | **Percentage** |
| --- | --- | --- |
| Yes | 111 | 19.86% |
| No | 239 | 42.75% |
| I am not sure | 209 | 37.39% |
| TOTAL | 559 | 100.00% |
| **How much education/training do researchers need to effectively use GenAI chatbots in scientific processes?** | **Responses** | **Percentage** |
| A lot | 203 | 39.42% |
| Some | 230 | 44.66% |
| Very little | 40 | 7.77% |
| None | 11 | 2.14% |
| I am not sure | 31 | 6.02% |
| TOTAL | 515 | 100.00% |
| **Are you interested in receiving education/training on GenAI chatbot use in scientific processes?** | **Responses** | **Percentage** |
| Yes | 369 | 72.07% |
| No | 48 | 9.38% |
| Maybe | 95 | 18.55% |
| TOTAL | 512 | 100.00% |
| **How important will GenAI chatbots be in future scientific research?** | **Responses** | **Percentage** |
| Very important | 280 | 54.37% |
| Important | 177 | 34.37% |
| Unimportant | 18 | 3.50% |
| Very unimportant | 2 | 0.39% |
| I'm not sure | 38 | 7.38% |
| TOTAL | 515 | 100.00% |
| **How will GenAI chatbots impact future scientific research?** | **Responses** | **Percentage** |
| Very positively | 130 | 25.19% |
| Positively | 250 | 48.45% |
| Negatively | 45 | 8.72% |
| Very negatively | 21 | 4.07% |
| I am not sure | 70 | 13.57% |
| TOTAL | 516 | 100.00% |

<sup>a</sup> Participants could choose more than one option.

A large proportion of respondents reported that their research institution did not offer guidance or training on GenAI tools (43.67%, 245/561), while fewer (20.50%, 115/561) indicated that their institution offered such training, but they had not yet taken it. Only 16.4% (92/561) reported having received training on GenAI chatbot use. In response to questions about institutional policies on GenAI chatbot use within the scientific process, 19.86% (111/559) indicated that their institution had implemented such policies. Yet, most respondents expressed a need for some (44.66%, 230/515) or a lot (39.42%, 203/515) of additional training and education to be necessary to effectively use GenAI chatbots in the scientific process. Additionally, a majority of respondents reported interest in receiving training on GenAI chatbot use in the scientific process (72.07%, 369/512).

Using a five-point scale ranging from very helpful to very unhelpful, respondents rated how useful they believed GenAI chatbots would be during different stages of the scientific process (see **Figure 1**). Notably, almost one third of respondents (31.91%, 164/514) perceived GenAI chatbots to be very helpful in conducting literature searches or reviews. Additionally, 31.76% (162/510) rated GenAI chatbots as very helpful for translating research materials, while 29.16% (149/511) rated them as very helpful for manuscript writing and editing. However, 20.31% (104/512) perceived GenAI chatbots to be unhelpful for generating presentations. For other evaluative tasks, such as peer review and research engagement, there were higher proportions of uncertainty and negative perceptions. Respondents reported being uncertain regarding the helpfulness of GenAI chatbots in peer reviewing and critiquing other researchers’ work (28.32%, 145/512) and increasing research engagement (28.04%, 143/512). Similarly, 22.66% (116/512) indicated they were unsure about how useful GenAI chatbots would be for analysing and interpreting data.

**Figure 1:**
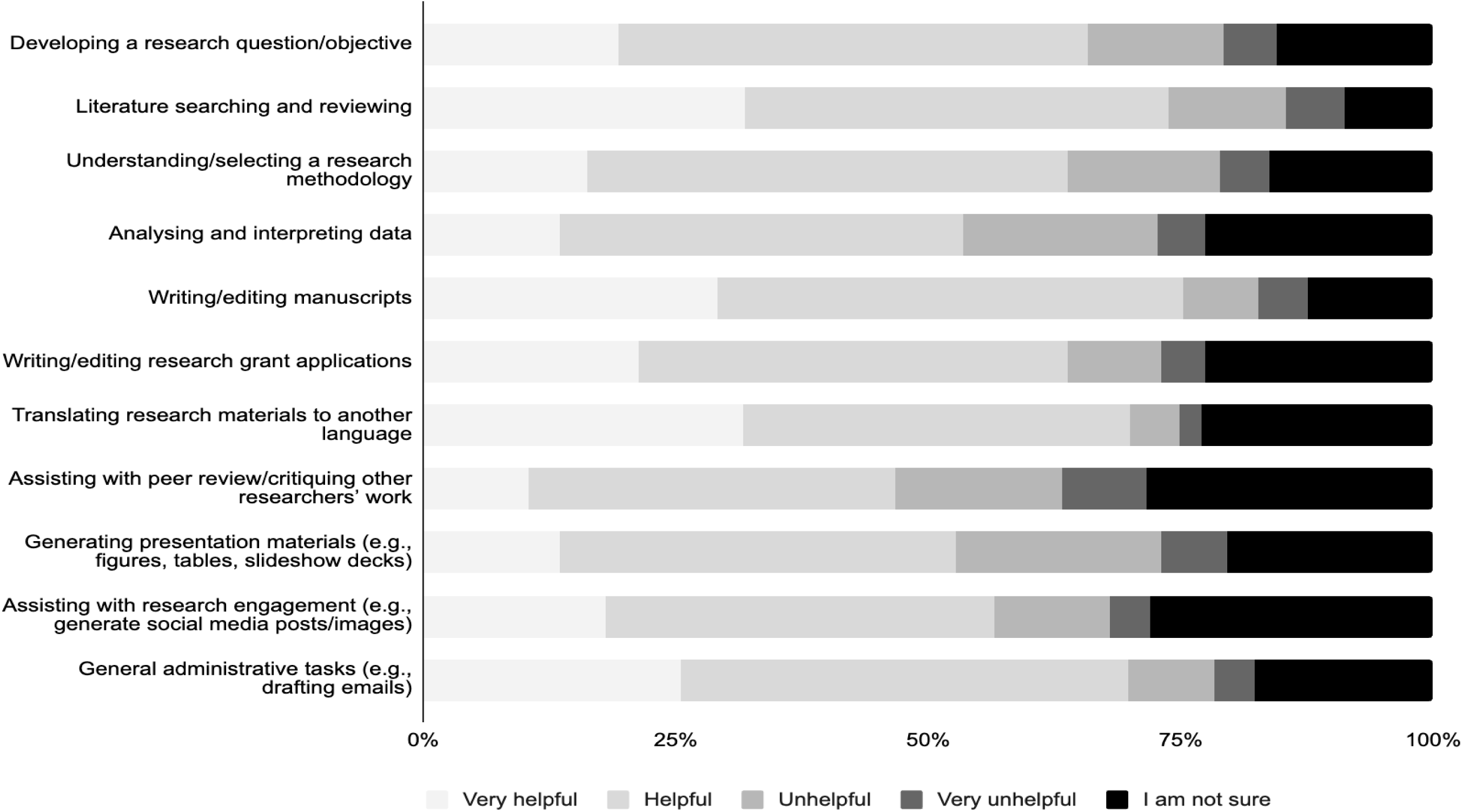
TCIM Researchers’ Perceptions of Steps in the Scientific Process Where GenAI Chatbots Are Helpful.

Researchers were also asked to rate their personal use of GenAI chatbots in the scientific process on a five-point scale with options ranging from always to never (see **Figure 2**). Respondents reported using GenAI chatbots most frequently to support writing-related tasks, with manuscript writing and editing having the highest regular use: 12.09% (62/511) reported always using GenAI chatbots for assistance, and 24.76% (127/511) reported using it often. Translation of research materials demonstrated a similar usage frequency, with 13.67% (70/512) using GenAI chatbots always and 22.46% (115/512) using it often. Other activities, including peer review and research engagement, had lower levels of use. For peer review, 46.39% (237/511) reported never using GenAI chatbots, while 44.81% (229/511) reported never using it for research engagement. Data analysis and interpretation also showed limited use, with more than one-third of participants (37.30%, 191/512) indicating they never used GenAI chatbots for this purpose. Overall, respondents used GenAI chatbots for drafting, translation, and writing-related purposes, and least for critical or interpretive aspects of the research process.

**Figure 2:**
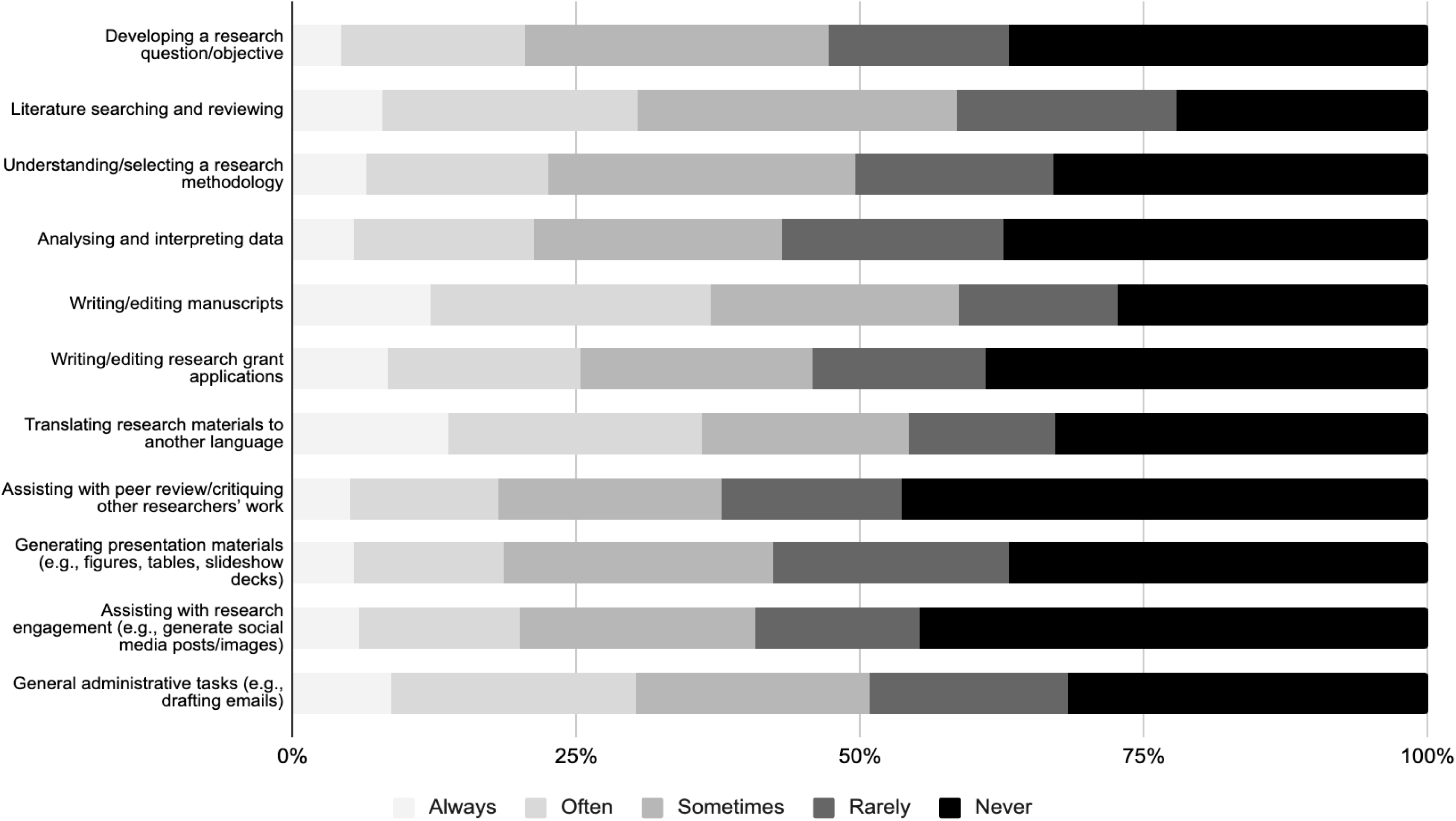
TCIM Researchers’ Frequency of GenAI Chatbot Use in Various Steps of the Scientific Process.

Respondents generally expressed strong agreement with many proposed benefits of GenAI chatbots in the scientific process, particularly those related to efficiency and accessibility (see **Figure 3**). On a five-point scale with options ranging from strongly disagree to strongly agree, the highest level of agreement (agree or strongly agree) among the respondents was observed for workload reduction (74.07%, 360/486) and increased efficiency in data analysis and experimentation (71.14%, 345/485). They also perceived benefits of GenAI chatbots in increasing the availability and accessibility of scientific information and assistance (70.37%, 342/486) and the handling and analysis of large datasets (70.10%, 340/485).

**Figure 3:**
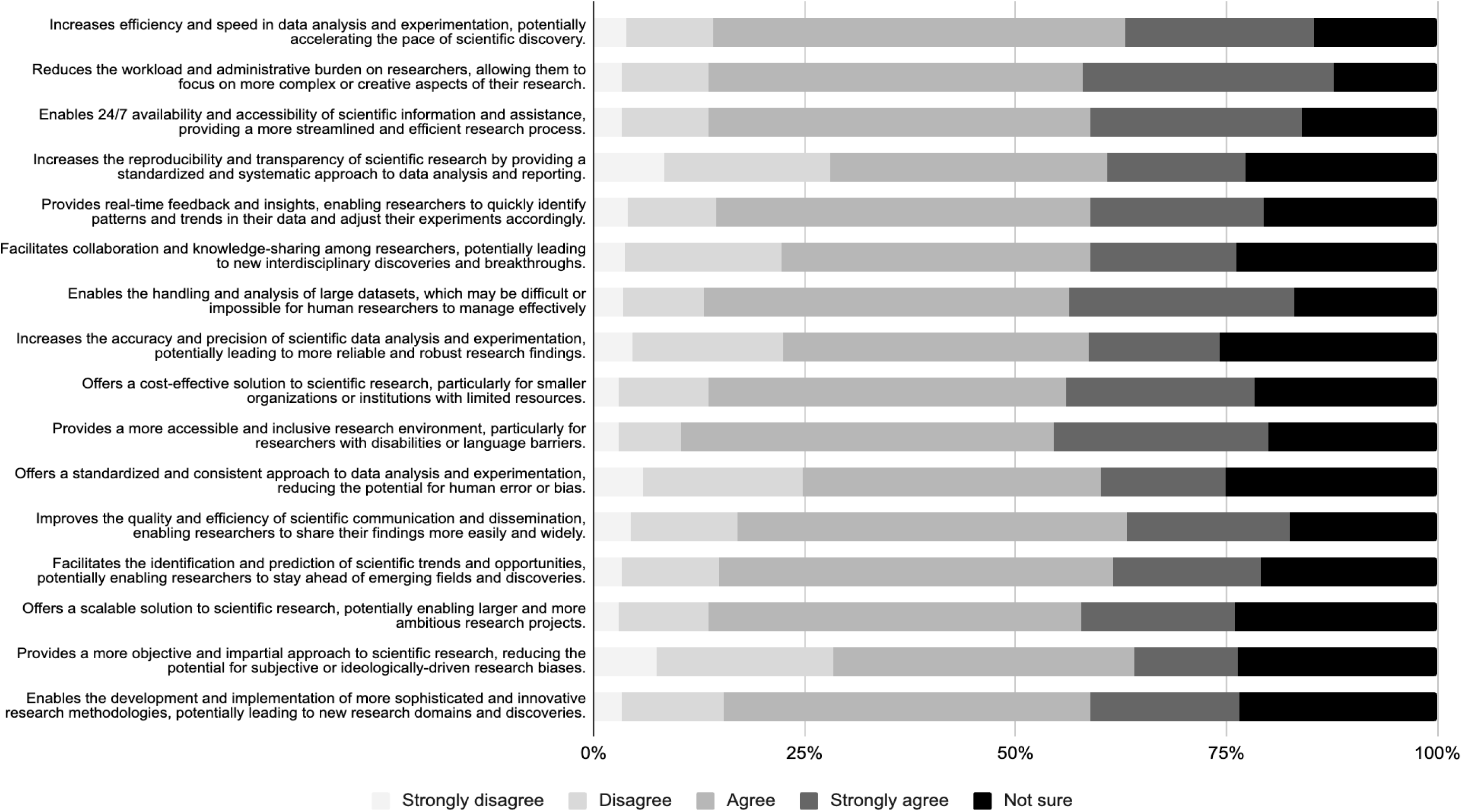
TCIM Researchers’ Perceptions of Proposed Benefits of GenAI Chatbots in the Scientific Process.

Using another five-point scale, with options ranging from strongly disagree to strongly agree, participants rated their attitudes regarding challenges associated with using GenAI chatbots in the scientific process (see **Figure 4**). Ethical and legal issues were perceived as significant challenges to GenAI chatbot use in the scientific process, with 80.17% (387/484) agreeing or strongly agreeing that GenAI chatbots raise concerns involving transparency, privacy, and consent. Concerns about transparency and interpretability were also prominent, with 72.41% (349/482) agreeing or strongly agreeing that limited transparency in decision-making is a major challenge. Other challenges included bias, errors, and limitations in understanding scientific complexity: 72.37% (351/485) agreed or strongly agreed that GenAI chatbots can produce biased or skewed data due to algorithmic limitations, while 66.73% (323/485) agreed that errors can arise from GenAI chatbots’ difficulty in interpreting scientific terminology.

**Figure 4:**
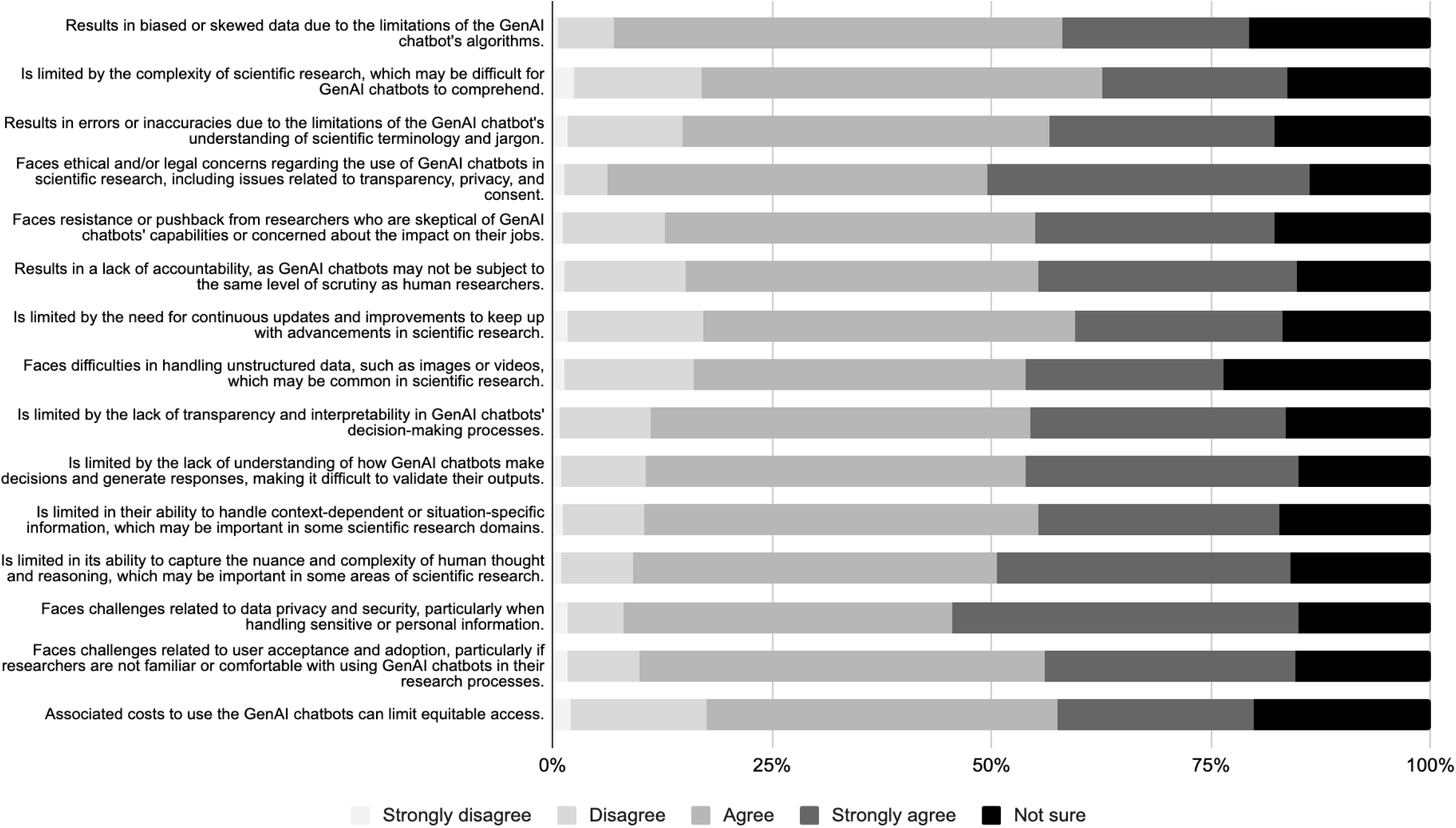
TCIM Researchers’ Perceptions of Proposed Challenges of GenAI Chatbots in the Scientific Process.

The open-ended questions provided respondents with an opportunity to elaborate on the types of GenAI chatbot-related training they had received and to reflect on additional benefits or challenges of GenAI chatbots in the scientific process beyond those listed in the survey. A complete summary of designated themes, subthemes, individual codes, and coding frequencies is available online, accompanied by full thematic analysis tallies and representative quotations (see https://osf.io/vezrf/files/ku58j).

Analysis of responses regarding GenAI chatbot training revealed several common themes, including the “Nature and Structure of GenAI Training”, “Integration of GenAI Training into Research Workflows”, “Ethical and Responsible Use of GenAI”, “Use of GenAI in Teaching and Educational Contexts”, and “Limited or Absent Institutional Training”. Respondents described experiences ranging from extensive multi-session or program-level training to informal awareness sessions or no structured training at all.

A parallel thematic analysis of additional perceived benefits identified themes, such as “Research Workflow Support”, “Communication and Accessibility Enhancements”, “Efficiency and Productivity Gains”, and a set of “Counterpoints and Caveats”, as well as a substantial number of respondents who reported no additional benefits beyond those already listed. The reported benefits were most related to literature synthesis, hypothesis generation, experimental design, translation support, public or lay communication of scientific information, and saving time in writing or administrative tasks. Others noted that GenAI chatbots could be used to provide a second review in various tasks, support idea development, or facilitate access to scientific resources.

The open-ended question on perceived challenges of GenAI chatbots in the scientific process resulted in a wide range of concerns that further contextualized the quantitative findings of this survey. The thematic analysis identified themes, such as the “Risk of Inaccuracy Hallucinations, and Unreliable Outputs”, “Bias, Transparency and Reproducibility Concerns”, “Environmental and Resource Costs”, “Impacts on Researcher Skill Development and Critical Thinking”, "Ethical, Legal, and Equity Implications”, and “Broader Structural Risks to the Scientific Ecosystem”.

Respondents highlighted issues such as fabricated citations, inconsistent or unverifiable claims, and the opacity of GenAI chatbot decision-making processes, raising concerns about erosion of research quality, traceability, and scientific rigour. Some respondents also emphasized environmental burdens, including high energy and water consumption, along with concerns that overreliance on GenAI chatbots can diminish researchers’ critical thinking and originality. Additional perceptions also suggested concerns regarding distinguishing authentic scholarship from AI-generated content.

## Discussion

The primary objective of this study was to examine TCIM researchers’ attitudes toward and perceptions of GenAI chatbot use across the scientific process. To our knowledge, this survey is the first to offer an international perspective on TCIM researchers’ familiarity with, use of, and views on GenAI chatbots, as well as their applications within the field. Overall, most respondents expressed positive views regarding the potential benefits of AI chatbots in the scientific process, while also acknowledging their disadvantages and challenges.

In our survey, most respondents reported familiarity with AI chatbots and prior use of these tools for research purposes, with ChatGPT being the most used. Respondents indicated that GenAI chatbots were used most frequently, and perceived as most helpful, for manuscript writing and editing, as well as for translating research materials, and generally viewed GenAI chatbots as having a positive impact on the future of scientific research. The most reported perceived benefits of GenAI chatbot use in the scientific process included increased efficiency in data analysis and experimentation, improved accessibility to scientific information and research assistance, enhanced handling and analysis of large datasets, and overall workload reduction. Similarly, when investigating the perceptions of medical researchers, the benefits of GenAI chatbots most frequently reported included reducing researchers’ workloads and administrative burden (66.9%), enabling constant availability and accessibility to scientific information and assistance (55.7%), improving the handling and analysis of large datasets compared with human researchers (56.1%), and increasing the efficiency and speed of data analysis (52.9%), among others [6].

Additionally, recent research on AI use in scientific processes has reported similar findings [3,33-35]. For example, a survey of 1,600 researchers found that AI tools were expected to improve data processing, accelerate computations, automate data acquisition, and reduce the time and resources required for research [36].

Despite the reported benefits and frequency of use, relatively few respondents indicated that their institutions offered training on the appropriate use of AI tools or chatbots in the scientific process, and an even smaller proportion reported the presence of formal institutional policies addressing their use. Given that most respondents expressed a desire to learn more about and receive training on GenAI chatbot use, this gap may reflect the recent adoption of GenAI chatbots in research and the restrictions on the use of AI tools in research articles imposed by journals [6,37].

Nonetheless, the absence of institutional training and policies raises important concerns regarding appropriate research application, scientific integrity, and ethical use. The most frequently reported challenges included the production of biased outputs, errors arising from GenAI’s limited ability to accurately interpret scientific terminology, and a lack of transparency and interpretability in AI-assisted decision-making. Consistent with these concerns, respondents reported the lowest use of GenAI chatbots for evaluative tasks, such as peer review and critical engagement with other researchers’ work. Additionally, ethical and legal considerations of GenAI chatbot use in the scientific process were also identified. Notably, 80.17% of respondents agreed or strongly agreed that GenAI chatbots raise concerns related to transparency, privacy, and consent. A previous cross-sectional survey of medical researchers [6] reported similar concerns regarding ethical and legal issues (76.8%), limited capacity of AI to capture the nuance and complexity of human reasoning (76.4%), lack of transparency and interpretability in decision-making processes (75.7%), data privacy and security risks (72.9%), and biased or skewed outputs (71.9%).

While many journals have taken steps to mitigate the challenges with AI use, such as requiring authors to disclose AI use, evidence suggests that undeclared ChatGPT-assisted manuscripts continue to be published in peer-reviewed journals [37,38]. Recent reports have also documented the retraction or correction of numerous published papers due to AI-generated inaccurate information, including fabricated references [39]. The development of clear guidelines and standardized training on the use and adoption of GenAI chatbots in research may help mitigate these risks [3,34,35], consistent with the views of many respondents in this study who reported a need for more education and training on the use of GenAI chatbots in the scientific process.

Ultimately, TCIM researchers hold distinct perspectives compared to those in biomedical fields due to the diversity of TCIM therapies, their frequent engagement with traditional knowledge systems, and the cultural nature of many TCIM approaches to patient care [15,40,41]. Consequently, they have differing views on the use of GenAI chatbots for study ideation, methodological design, and data sovereignty. By exploring these attitudes and perceptions, the study identified the perceived benefits, challenges, and ethical considerations associated with integrating GenAI tools into TCIM research workflows. The findings may have important implications for the responsible adoption of GenAI chatbots in TCIM research, providing valuable insights into researchers’ acceptance of these tools and the potential barriers to their effective use. Understanding TCIM researchers’ perspectives may also inform the development of GenAI technologies that align more closely with the specific needs and values of the TCIM research community, ultimately supporting the advancement of high-quality, evidence-based research in this field [42]. Furthermore, this study will contribute to broader discussions about the ethical use of GenAI chatbots in scientific research, emphasizing the importance of transparency, accountability, and equity in the design and implementation of these technologies [5]. Outside of the direct research context, our study findings may also inform how TCIM institutions, journals, and professional networks develop policies and guidance to support the responsible and ethical use of GenAI chatbots in research workflows.

### Implications for TCIM Research Practice

GenAI tools have the potential to meaningfully strengthen TCIM research practice by supporting interdisciplinary collaboration and improving methodological coherence across diverse evidence traditions. Within integrative medicine teams, where clinicians and researchers often navigate differing terminologies, epistemologies, and documentation norms, GenAI chatbots can facilitate communication by synthesizing information across systems, translating concepts between TCIM and conventional medicine frameworks, and helping harmonize documentation and reporting standards used in clinical trials. These capabilities may also advance transparency and rigour in TCIM research by assisting with standardized data extraction, protocol adherence checks, and consistent reporting structures that align with international guidelines. Additionally, GenAI tools can expand the capacity for meta-research in TCIM by enabling efficient mapping, appraisal, and synthesis of heterogeneous evidence bases, thereby supporting more robust evaluations of research quality and methodological trends. Together, these applications underscore the role of GenAI technology as a potential catalyst for improving integrative research environments and strengthening the evidence foundations underpinning TCIM practice.

### Strengths and Limitations

Our study had several notable strengths. Conducting a cross-sectional survey enabled the collection of data at a single time point, which made the study efficient and cost-effective [43]. By employing a sampling strategy that sought a large and diverse international participant pool, we enhanced the generalizability of our findings regarding TCIM researchers’ attitudes and perceptions of GenAI chatbots in the scientific process. Additionally, all data extraction and cleaning procedures were conducted first by software, then manually verified, ensuring accuracy and minimizing errors.

However, the study also had limitations [44]. Despite planning to survey many researchers, we anticipated a low response rate due to several factors. These included participants unfamiliar with the research team and not expecting a survey invitation. Other logistical challenges, such as outdated contact information, changes in affiliations, email inaccessibility, bounce-back emails, vacations, and retirements, may have also contributed to the nonresponse bias. Furthermore, non-English-speaking researchers were largely excluded from the sample due to language and resource constraints. This may have had implications for TCIM knowledge representation, as non-English speaking researchers working with non-biomedical, spiritual, symbolic, or narrative knowledge systems prevalent in TCIM could have been underrepresented, limiting the diversity of epistemic perspectives. Limited English proficiency among respondents may have also affected the completion of the survey, potentially restricting a comprehensive list of attitudes and opinions. Consequently, this study may not have been applicable to researchers who published in languages other than English. In the context of recent developments in large language models capable of automated multilingual translation, future research should employ multilingual survey tools combined with AI-based translation scripts to ensure global participation can be accommodated. Furthermore, accessibility restrictions in certain regions may have introduced selection bias. Although participants were allowed to share their experiences with any type of GenAI chatbot (e.g., ChatGPT, Google Gemini, Ernie Bot, DeepSeek), the use of a Western survey platform, along with regional differences in access to GenAI tools, may have limited participation from other world regions. These findings may also have been skewed towards those who were interested in or had experience using GenAI chatbots, as researchers who were skeptical or less engaged with these technologies may have been less inclined to participate. Another limitation could have been recall bias; participants may have had difficulty accurately remembering their use of GenAI chatbots or may have unintentionally omitted details about their experiences, especially if significant time had elapsed since the research activities that were being surveyed took place. Finally, the survey primarily focused on GenAI chatbots rather than rule-based or retrieval-based systems, which may have limited its applicability to the full spectrum of AI tools used in TCIM.

## Conclusion

This study offers insights into how TCIM researchers perceive the role of GenAI chatbots in the scientific process. By exploring their attitudes, experiences, and ethical concerns, the findings may help guide the responsible development and implementation of these tools in TCIM research. The results may also inform efforts to enhance research quality, support interdisciplinary collaboration, and promote equitable access to GenAI technologies. Ultimately, it is hoped that this work will contribute to ongoing discussions about evidence-based practices in TCIM and the thoughtful integration of GenAI chatbots across diverse research contexts.

## List of Abbreviations

AI: artificial intelligence
CHERRIES: checklist for reporting results of internet e-surveys
DOI: digital object identifier
GenAI: generative artificial intelligence
NCCIH: National Center for Complementary and Integrative Health
OSF: Open Science Framework
PMID: PubMed Identifier
STROBE: Strengthening the Reporting of Observational Studies in Epidemiology
TCIM: traditional, complementary and integrative medicine
WHO: World Health Organization

## Declarations

### Ethics Approval and Consent to Participate

Ethics approval was sought and obtained from the University Hospital Tübingen Research Ethics Board (REB #: 079/2025BO2) to conduct this study.

### Consent for Publication

All authors consent to this manuscript’s publication.

### Availability of Study Materials and Data

All relevant study materials and data are included in this manuscript or posted on the Open Science Framework at https://doi.org/10.17605/OSF.IO/VEZRF.

### Authors’ Contributions

JYN: designed and conceptualized the study, collected and analysed data, co-drafted the manuscript, and gave final approval of the version to be published.

JT: collected and analysed data, co-drafted the manuscript, and gave final approval of the version to be published.

NS: analysed data, made critical revisions to the manuscript, and gave final approval of the version to be published.

KA: assisted with the collection and analysis of data, made critical revisions to the manuscript, and gave final approval of the version to be published.

PKG: assisted with the collection and analysis of data, made critical revisions to the manuscript, and gave final approval of the version to be published.

SL: assisted with the collection and analysis of data, made critical revisions to the manuscript, and gave final approval of the version to be published.

DM: assisted with the collection and analysis of data, made critical revisions to the manuscript, and gave final approval of the version to be published.

MR: assisted with the collection and analysis of data, made critical revisions to the manuscript, and gave final approval of the version to be published.

MS: assisted with the collection and analysis of data, made critical revisions to the manuscript, and gave final approval of the version to be published.

JPS: assisted with the collection and analysis of data, made critical revisions to the manuscript, and gave final approval of the version to be published.

TY: assisted with the collection and analysis of data, made critical revisions to the manuscript, and gave final approval of the version to be published.

MSL: assisted with the collection and analysis of data, made critical revisions to the manuscript, and gave final approval of the version to be published.

HC: assisted with the collection and analysis of data, made critical revisions to the manuscript, and gave final approval of the version to be published.

## References

1. Xu Y, Liu X, Cao X, Huang C, Liu E, Qian S, Liu X, Wu Y, Dong F, Qiu CW, Qiu J, Hua K, Su W, Wu J, Xu H, Han Y, Fu C, Yin Z, Liu M, Roepman R, Dietmann S, Virta M, Kengara F, Zhang Z, Zhang L, Zhao T, Dai J, Yang J, Lan L, Luo M, Liu Z, An T, Zhang B, He X, Cong S, Liu X, Zhang W, Lewis JP, Tiedje JM, Wang Q, An Z, Wang F, Zhang L, Huang T, Lu C, Cai Z, Wang F, Zhang J. Artificial intelligence: A powerful paradigm for scientific research. Innov. 2021;2(4):100179. 10.1016/j.xinn.2021.100179

2. Zhang P, Zhang Q, Li S. Advancing cancer prevention through an AI-based integration of traditional and Western medicine. Cancer Discov. 2024;14(11):2033–2036. 10.1158/2159-8290.CD-24-0832

3. Salvagno M, Taccone FS, Gerli AG. Can artificial intelligence help for scientific writing? Crit Care. 2023;27(1). 10.1186/s13054-023-04380-2

4. Chen A, Liu L, Zhu T. Advancing the democratization of generative artificial intelligence in healthcare: A narrative review. J Hosp Manag Health Policy. 2024;8:12. 10.21037/jhmhp-24-54

5. Singhal A, Neveditsin N, Tanveer H, Mago V. Toward fairness, accountability, transparency, and ethics in AI for social media and health care: Scoping review. JMIR Med Inform. 2024;12(1):e50048. 10.2196/50048

6. Ng JY, Maduranayagam SG, Suthakar N, Li A, Lokker C, Iorio A, Haynes RB, Moher D. Attitudes and perceptions of medical researchers towards the use of artificial intelligence chatbots in the scientific process: An international cross-sectional survey. Lancet Digit Health. 2025;7(1):e94–102. 10.1016/S2589-7500(24)00202-4

7. Fabiano N, Gupta A, Bhambra N, Luu B, Wong S, Maaz M, Fiedorowicz JG, Smith AL, Solmi M. How to optimize the systematic review process using AI tools. JCPP Adv. 2024;4(2). 10.1002/jcv2.12234

8. Kirchner GJ, Kim RY, Weddle J, Bible JE. Can artificial intelligence improve the readability of patient education materials? Clin Orthop Relat Res. 2023. 10.1097/corr.0000000000002668

9. Ng JY, Dhawan T, Dogadova E, Taghi-Zada Z, Vacca A, Wieland LS, Moher D. Operational definition of complementary, alternative, and integrative medicine derived from a systematic search. BMC Complement Med Ther. 2022;22:104. 10.1186/s12906-022-03556-7

10. World Health Organization. Traditional medicine [Internet]. WHO; 2023 [cited 2025 Jun 25]. Available from: https://www.who.int/news-room/questions-and-answers/item/traditional-medicine

11. World Health Organization. Traditional, complementary and integrative medicine [Internet]. WHO; 2019 [cited 2025 Jun 25]. Available from: https://www.who.int/health-topics/traditional-complementary-and-integrative-medicine#tab=tab_1

12. National Center for Complementary and Integrative Health. Complementary, alternative, or integrative health: What’s in a name? [Internet]. NCCIH; 2021 [cited 2025 Jun 25]. Available from: https://www.nccih.nih.gov/health/complementary-alternative-or-integrative-health-whats-in-a-name

13. Ng JY, Boon HS, Thompson AK, Whitehead CR. Making sense of “alternative”, “complementary”, “unconventional” and “integrative” medicine: Exploring the terms and meanings through a textual analysis. BMC Complement Altern Med. 2016;16(1):134. 10.1186/s12906-016-1111-3

14. Raja M, Cramer H, Lee MS, Wieland LS, Ng JY. Addressing the challenges of traditional, complementary, and integrative medicine research: An international perspective and proposed strategies moving forward. Perspect Integr Med. 2024;3(2):86–97. 10.56986/pim.2024.06.004

15. Veziari Y, Leach MJ, Kumar S. Barriers to the conduct and application of research in complementary and alternative medicine: A systematic review. BMC Complement Altern Med. 2017;17(1):166. 10.1186/s12906-017-1660-0

16. Ng JY. Insight into the characteristics of research published in traditional, complementary, alternative, and integrative medicine journals: A bibliometric analysis. BMC Complement Med Ther. 2021;21(1). 10.1186/s12906-021-03354-7

17. World Health Organization. WHO traditional medicine strategy: 2014-2023 [Internet]. Geneva: World Health Organization; 2013 [cited 2025 Aug 14]. Available from: https://www.who.int/publications/i/item/9789241506096.

18. Ng JY, Cramer H, Lee MS, Moher D. Traditional, complementary, and integrative medicine and artificial intelligence: Novel opportunities in healthcare. Integr Med Res. 2024;13(1):101024. 10.1016/j.imr.2024.101024

19. Amabie T, Izah SC, Ogwu MC, Hait M. Harmonizing tradition and technology: The synergy of artificial intelligence in traditional medicine. In: Herbal Medicine Phytochemistry: Applications and Trends 2023 Nov 25 (pp. 1-23). Cham: Springer International Publishing. 10.1007/978-3-031-21973-3_70-1

20. Ofosu-Asare Y. Cognitive imperialism in artificial intelligence: Counteracting bias with indigenous epistemologies. AI & Soc. 2024:1–7. 10.1007/s00146-024-02065-0

21. World Health Organization. Mapping the application of artificial intelligence in traditional medicine: Technical brief [Internet]. WHO; 2025 [cited 2025 Jun 25]. Available from: https://www.who.int/publications/i/item/9789240107663

22. Elendu C, Amaechi DC, Elendu TC, Jingwa KA, Okoye OK, Okah MJ, Ladele JA, Farah AH, Alimi HA. Ethical implications of AI and robotics in healthcare: A review. Medicine. 2023;102(50):e36671. 10.1097/MD.0000000000036671

23. Ng JY, Tan J, Adapa K, Gupta PK, Li S, Mehta DH, Ring M, Shridhar M, Souza JP, Yoshino T, Lee MS, Cramer H. Traditional, complementary, and integrative medicine researcher attitudes and perceptions of generative artificial intelligence chatbots in the scientific process: A large-scale, international cross-sectional survey [Internet]. OSF; 2025 [cited 2025 Jun 25]. Available from: 10.17605/OSF.IO/Q9UNK

24. von Elm E, Altman DG, Egger M, Pocock SJ, Gøtzsche PC, Vandenbroucke JP. Strengthening the Reporting of OBservational studies in Epidemiology (STROBE) statement: Guidelines for reporting observational studies. BMJ. 2007;335(7624):806–8. 10.1136/bmj.39335.541782.ad

25. Eysenbach G. Improving the quality of web surveys: The CHEcklist for Reporting Results of Internet E-Surveys (CHERRIES). J Med Internet Res. 2004;6(3):e34. 10.2196/jmir.6.3.e34

26. Fantini D. easyPubMed: Search and Retrieve Scientific Publication Records from PubMed [Internet]. R-Packages; 2019 [cited 2025 Jun 25]. Available from: https://cran.r-project.org/web/packages/easyPubMed/index.html

27. Fry D, Al-Khafaji, W. RedEye Pipeline [Software]. (V1.0). Toronto: Daniel Fry. (2025). [Accessed 2025 Aug 29]. Retrieved from: 10.5281/zenodo.16996504

28. SurveyMonkey Inc. SurveyMonkey [Internet]. San Mateo: SurveyMonkey Inc.; 2026 [cited 2025 Aug 14]. Available from: https://www.surveymonkey.com/.

29. Ng JY, Santoro LJ, Cobey KD, Steel A, Cramer H, Moher D. Complementary, alternative, and integrative medicine researchers’ practices and perceived barriers related to open science: An international, cross-sectional survey. PloS ONE. 2024;19(5):e0301251–1. 10.1371/journal.pone.0301251

30. Ng JY, Lin BX, Kreuder L, Cramer H, Moher D. Open science practices among authors published in complementary, alternative, and integrative medicine journals: An international, cross-sectional survey. Medicine. 2024;103(44):e40259. 10.1097/MD.0000000000040259

31. Raskind IG, Shelton RC, Comeau DL, Cooper HLF, Griffith DM, Kegler MC. A review of qualitative data analysis practices in health education and health behavior research. Health Educ Behav. 2019;46(1):32–9. 10.1177/1090198118795019

32. Thomas DR. A general inductive approach for analyzing qualitative evaluation data. Am J Eval. 2006;27(2):237–46. 10.1177/1098214005283748

33. Noy S, Zhang W. Experimental evidence on the productivity effects of generative artificial intelligence. Science. 2023;381(6654):187–192. 10.1126/science.adh2586.

34. Salvagno M, Taccone FS, Gerli AG. Can artificial intelligence help for scientific writing? Crit Care. 2023;27(1):75. 10.1186/s13054-023-04380-2.

35. Khalifa M, Albadawy M. Using artificial intelligence in academic writing and research: An essential productivity tool. Comput Methods Programs Biomed Update. 2024;5:100145. 10.1016/j.cmpbup.2024.100145.

36. Van Noorden R, Perkel JM. AI and science: What 1,600 researchers think. Nature. 2023;621(7980):672–675. 10.1038/d41586-023-02980-0.

37. Brainard J. Journals take up arms against AI-written text. Science. 2023;379(6634):740–741. 10.1126/science.adh2762.

38. Lund, B.D. and Naheem, K.T. (2024), Can ChatGPT be an author? A study of artificial intelligence authorship policies in top academic journals. Learned Publishing. 2023;37(1):13-21. 10.1002/leap.1582

39. Chen A, Chen DO. Accuracy of chatbots in citing journal articles. JAMA Netw Open. 2023;6(8):e2327647. 10.1001/jamanetworkopen.2023.27647

40. Ng JY, Dhawan T, Fajardo RG, Masood HA, Sunderji S, Wieland LS, Moher D. The brief history of complementary, alternative, and integrative medicine terminology and the development and creation of an operational definition. Integr Med Res. 2023;12(4):100978. 10.1016/j.imr.2023.100978

41. Ng JY, Lee MS, Liu JP, Steel A, Wieland LS, Witt CM, Moher D, Cramer H. How can meta-research be used to evaluate and improve the quality of research in the field of traditional, complementary, and integrative medicine?. Integr Med Res. 2024;13(3):101068. 10.1016/j.imr.2024.101068

42. Wang L, Tang K, Zhang P, Liu J, Wu B, Chen J, Li Y, Du S, Wang Y, Li S. Development and validation of an interpretable machine learning model for non-invasive screening of precancerous gastric lesions using symptom and lifestyle data: A multicentre cohort study. eClinicalMedicine. 2026;92:103756. 10.1016/j.eclinm.2026.103756

43. Wang X, Cheng Z. Cross-sectional studies: Strengths, weaknesses, and recommendations. Chest. 2020;158(1):65–71. 10.1016/j.chest.2020.03.012

44. Choi BC, Pak AW. A catalog of biases in questionnaires. Prev Chronic Dis. 2004;2(1):A13. https://www.cdc.gov/pcd/issues/2005/jan/04_0050.htm

